# Utilization of Whole Genome Sequencing to Understand SARS-CoV-2 Transmission Dynamics in Long-Term Care Facilities, Correctional Facilities and Meat Processing Plants in Minnesota, March – June 2020

**DOI:** 10.1101/2020.12.30.20248277

**Authors:** Nicholas B Lehnertz, Xiong Wang, Jacob Garfin, Joanne Taylor, Jennifer Zipprich, Brittany VonBank, Karen Martin, Dana Eikmeier, Carlota Medus, Brooke Wiedinmyer, Carmen Bernu, Matthew Plumb, Kelly Pung, Margaret A. Honein, Rosalind Carter, Duncan MacCannell, Kirk E. Smith, Kathryn Como-Sabetti, Kris Ehresmann, Richard Danila, Ruth Lynfield

## Abstract

Congregate settings and high-density workplaces have endured a disproportionate impact from COVID-19. In order to provide further understanding of the transmission patterns of SARS-CoV-2 in these settings, whole genome sequencing (WGS) was performed on samples obtained from 8 selected outbreaks in Minnesota from March – June, 2020. WGS and phylogenetic analysis was conducted on 319 samples, constituting 14.4% of the 2,222 total SARS-CoV-2-positive individuals associated with these outbreaks. Among the sequenced specimens, three LTCFs and both correctional facilities had spread associated with a single genetic sequence. A fourth LTCF had outbreak cases associated with two distinct sequences. In contrast, cases associated with outbreaks in the two meat processing plants represented multiple SARS-CoV-2 sequences. These results suggest that a single introduction of SARS-CoV-2 into a facility can result in a widespread outbreak, and early identification and cohorting of cases, along with continued vigilance with infection prevention and control measures is imperative.

## Introduction

COVID-19 has disproportionately impacted adults residing in long-term care facilities (LTCFs) in the United States (*1-5*). LTCF outbreaks have caused high numbers of hospitalizations and death. Similar findings have been reported in correctional facilities, with SARS-CoV-2 infection incidence among inmates and staff over five times greater and age-adjusted mortality over three times greater than that of the general population (*6-8*). Workers in high-density workplaces (e.g., meat processing plants) have similarly been heavily affected, with a disproportionate impact on minority populations (*9-11*).

The first identified COVID-19 case in Minnesota was detected on March 6, 2020. Shortly thereafter, COVID-19 outbreaks occurred across the state, including in LTCFs (March 12, 2020) and meat processing plants (March 15, 2020), followed shortly thereafter by correctional facilities (March 25, 2020). From March 6 through June 30, 2020, the Minnesota Department of Health (MDH) identified and responded to 1,060 distinct outbreaks of COVID-19 in LTCFs, comprising 4,421 resident cases and 3,002 staff cases. In addition, there were 4 discrete outbreaks in correctional facilities that resulted in a total of 382 cases, and 68 outbreaks in meat processing plants resulting in approximately 2,616 cases among employees^1^; outbreaks in these 3 settings accounted for 31.3% of all identified cases in Minnesota.

For outbreaks in both congregate settings and high-density workplaces, it was difficult to confirm the temporal and relational aspect of SARS-CoV-2 transmission; the role of intra-facility spread versus multiple introductions was difficult to disentangle. Whole-genome sequencing (WGS) of outbreak case specimens can be used to understand transmission dynamics and relatedness of viral pathogens in infectious disease outbreaks (*12-15*). Unprecedented efforts to sequence SARS-CoV-2 genomes have occurred at the local, regional, national and international level in order to investigate potential reinfections (*16-19*), nosocomial transmission (*20*), patterns of community spread (*21-23*), and sources of introduction of SARS-CoV-2 without known epidemiologic links (*24*).

In Minnesota, as part of the CDC SARS-CoV-2 Sequencing for Public Health Emergency Response, Epidemiology and Surveillance (SPHERES) consortium, the Minnesota Molecular Surveillance of SARS-CoV-2 (MN-SOS) initiative solicited outbreak specimens for sequencing and genetic variation analysis to understand viral transmission patterns in congregate settings and meat processing plants. WGS of a convenience sample of SARS-CoV-2 positive specimens associated with outbreaks was performed to supplement epidemiological information and assess whether single or multiple introductions likely occurred during a facility outbreak, and to evaluate molecular relatedness.

## Methods

Three types of outbreak settings (LTCF, correctional facilities and meat processing plants) were chosen for WGS, with selection of specific facilities based in part on the impact and severity of the outbreak, the need for further clarity regarding transmission patterns, and the availability of samples. Selected outbreaks occurred between March 6 and June 30, 2020 at four unique LTCFs (LTCF A-D), two correctional facilities (Correctional Facilities A and B), and two meat processing plants (Processing Plant A and B), along with identified cases in individuals residing in the same county as Processing Plant A (Community Samples A). An outbreak at a LTCF was defined as ≥1 confirmed COVID-19 case in a resident or staff member. In correctional facilities, an outbreak was defined as 1) ≥2 cases in the inmate population >7 days after intake to a new facility, with an epidemiologic link^2^, or 2) ≥2 cases in correctional staff with an epidemiologic link^3^ at the facility, or 3) ≥1 facility-acquired^4^ COVID-19 cases in an inmate. An outbreak at meat processing facilities was defined as ≥3 laboratory-confirmed COVID-19 cases among workers at a facility who resided in separate households. On June 1, a time component was added to the outbreak definition in meat processing plants, adding that case onset dates are within 14 days of each other. Cases at all outbreak locations were defined as an individual testing SARS-CoV-2 positive by reverse transcription polymerase chain reaction (RT-PCR) using the original CDC assay protocol (*25*).

Epidemiologic data, including gender, age, symptom status, symptom onset date, residence, occupation, and potential source of exposure were collected via interview with persons having lab-confirmed SARS-CoV-2.

WGS was performed by the MDH Public Health Laboratory (PHL) on available SARS-CoV-2 RT-PCR positive specimens, collected from March 6 through June 30, 2020. Specimens were obtained from the nasopharynx, the anterior nares or oropharynx. SARS-CoV-2 RNA extracts were acquired either as residual from clinical testing at MDH-PHL or other clinical laboratories serving Minnesota residents. cDNA and tiled amplicons were created as described in the ARTIC Network nCoV-2019 sequencing protocol (*26*). Sequencing Libraries for Illumina next-generation sequencing were prepared according to the Nextera DNA Flex protocol created by the State Public Health Bioinformatics Group (StaPH-B) (*27*) and sequenced using 2×250 bp Illumina V2 chemistry on MiSeq instruments. Consensus SARS-CoV-2 genome sequences for each specimen were generated with the StaPH-B Toolkit Monroe pipeline (*28*). Assembled SARS-CoV-2 genomes were individually reviewed in Geneious Prime 2019.2.1 (*29*) and genomes with gaps greater than 125 nucleotides were discarded.^5^

The Augur toolkit (*30*) was used for alignment of SARS-CoV-2 genome consensus sequences, phylogenetic tree generation, and incorporation of epidemiological sequence metadata. Genomes were aligned with MAFFT version 7.310 with options “--keeplength -- reorder --anysymbol --nomemesave --adjustdirection” (*31*). Variation in sequences identified in the first 54 and last 67 bases of the Wuhan-Hu-1 (GenBank: MN908947.3) reference sequence was masked during tree generation due to the inability of the tiled-amplicon sequencing approach to reliably generate sequence in those regions. IQ-TREE version 1.6.1 was used for phylogenetic tree creation with parameters “-ninit 2 -n 2 -me 0.05” (*32*). Output from Augur was visualized using Auspice as hosted by the nextstrain team at http://auspice.us/ (*30*). The resulting trees were visualized with the Interactive Tree of Life (iTOL) (*33*) with branch lengths rounded and scaled represent mutations from the reference. Pangolin lineages for all samples were retrieved following submission of assemblies to GISAID (*30, 34-35*).

Genetically closely related sequences (i.e., clusters) were defined as cases which were associated with an outbreak and formed a monophyletic clade on the state-wide phylogenetic tree.

In accordance with federal human subjects protection regulations at 45 CFR §46.101c and §46.102d and with the Guidelines for Defining Public Health Research and Public Health Non-Research, the project was reviewed by a human subjects protection coordinator at the CDC and in Minnesota DOH and determined to be a non-research, public health response exempt from institutional review board evaluation.

## Results

WGS and subsequent phylogenetic analysis was successfully conducted on 319 samples associated with the eight outbreaks, constituting 14.4% of the 2,222 total positive cases identified from outbreaks in Minnesota through June 2020. Specimens were obtained from staff and residents from four different LTCFs (180 specimens sequenced out of 505 identified cases, 35.6%), staff and inmates from 2 different correctional facilities (110 specimens sequenced out of 544 identified cases, 20.2%) and among employees at 2 different meat processing plants, along with identified community cases (29 samples sequenced out of 1,173 identified cases, 2.5%) (Table 1). Among most of the specimens that were sequenced, three LTCFs and both correctional facilities had spread associated with a single genetic sequence unique to each outbreak facility. A fourth LTCF had outbreak cases associated with two distinct sequences. In contrast, cases associated with outbreaks in the two meat processing plants represented multiple SARS-CoV-2 sequences. (Figure 1)

**Table 1.** Features of outbreaks and convenience samples of specimens collected and characterized by whole genome sequencing at long-term care facilities (LTCFs), correctional facilities and meat processing plants in Minnesota: March 6 – June 30, 2020^6^

| Outbreak Facility | Outbreak Dates | Total Confirmed Outbreak Cases at Facility | Total Samples Successfully Sequenced from Facility<br>N (%) | Role at Facility | Total Confirmed Outbreak Cases at Facility by Role | Total Samples Successfully Sequenced by Role<br>N (%) |
| --- | --- | --- | --- | --- | --- | --- |
| LTCF A | April 15 – June 11 | 89 | 27 (30.3%) | Staff | 38 | 10 (26.3%) |
|  |  |  |  | Residents | 51 | 17 (33.3%) |
| LTCF B | April 29 – June 11 | 190 | 82 (43.2%) | Staff | 76 | 5 (6.6%) |
|  |  |  |  | Residents | 114 | 77 (67.5%) |
| LTCF C | April 24 – June 30 | 139 | 32 (23.0%) | Staff | 56 | 23 (41.0%) |
|  |  |  |  | Residents | 83 | 9 (10.8%) |
| LTCF D | April 17 – May 15 | 74 | 39 (52.7%) | Staff | 21 | 3 (14.2%) |
|  |  |  |  | Residents | 53 | 36 (67.9%) |
| Correctional Facility A | March 25 – July 20 | 128 | 49 (38.3%) | Staff | 82 | 15 (18.3%) |
|  |  |  |  | Inmates | 46 | 34 (73.9%) |
| Correctional Facility B | May 13 – August 5 | 416 | 61 (14.7%) | Staff | 210 | 1 (0.5%) |
|  |  |  |  | Inmates | 206 | 60 (29.1%) |
| Processing Plant A | March 15 – Ongoing | 432 | 16 (3.7%) | Employees | 432 | 16 (3.7%) |
| Processing Plant B | April 11 – Ongoing | 724 | 5 (0.7%) | Employees | 724 | 5 (0.7%) |
| Community Samples A | May 15 – June 1 | 17 | 8 (47.1%) | Known Contact | 9 | 2 (22.2%) |
|  |  |  |  | No Known Contact | 8 | 6 (75.0%) |
| TOTALS |  | 2,222 | 319 (14.4%) |  |  |  |

**Figure 1.**
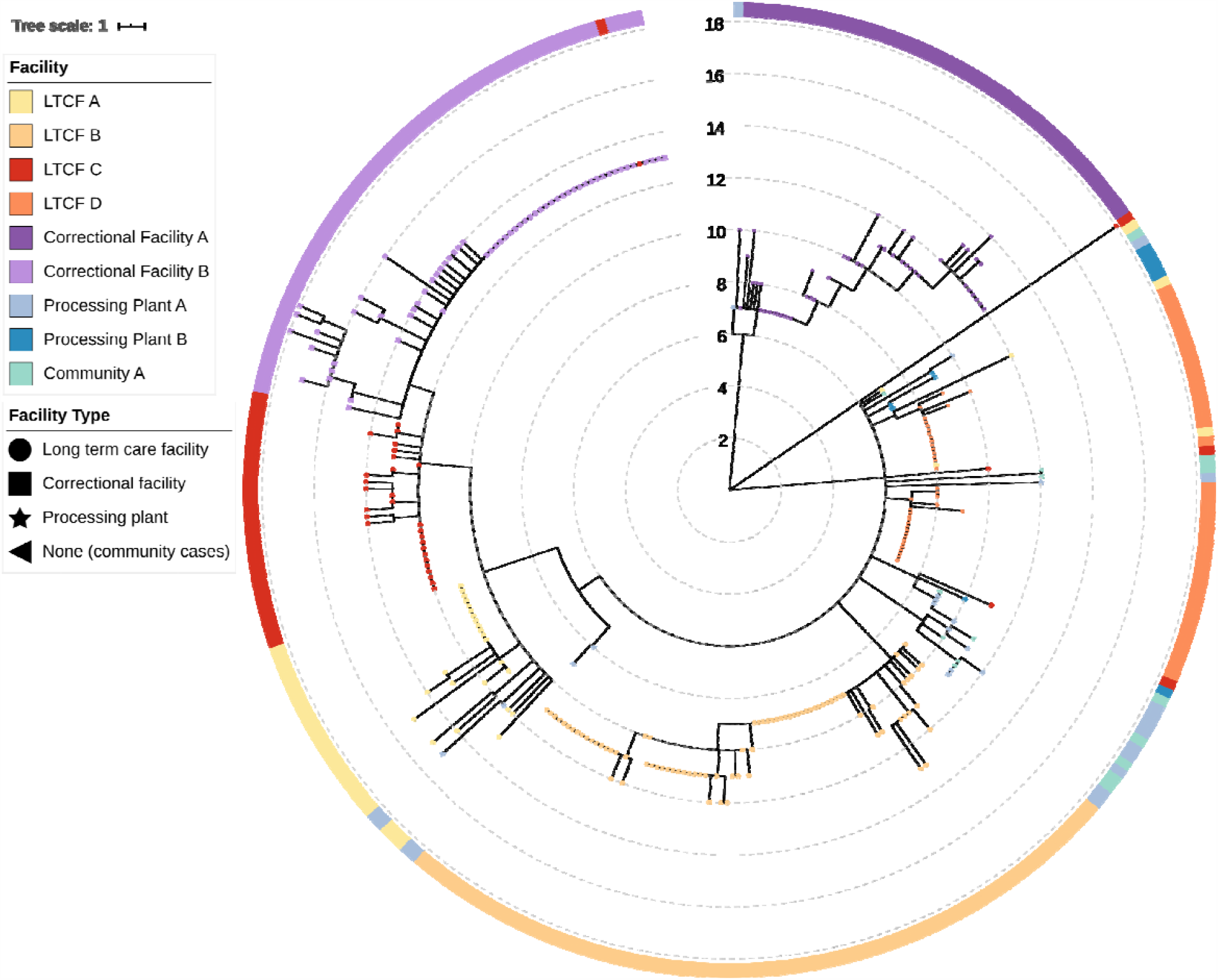
Phylogenetic tree of SARS-CoV-2 viruses associated with selected outbreaks in Minnesota, March 6 – June 30, 2020. IQ-TREE was used with the GTR substitution model for tree generation. Branch lengths were scaled to represent number of single nucleotide mutations as shown in the scale key.

### Single Cluster in Long-Term Care Facilities

During the COVID-19 outbreak at LTCF A^7^ between April 15 and June 11 (Figure 2A), 51 of 77 residents, and 38 of 108 health care personnel (HCP) tested were confirmed to be infected, following identification of positive HCP. Specimens from 17 residents (33.3% of cases) and 10 staff (26.3% of cases) were available for WGS. SARS-CoV-2 viral sequences from these 27 persons were genetically closely related (Pangolin lineage B.1.2). Viral genomes from two HCP sampled on April 30 and one resident sampled on May 18 at facility A did not cluster with each other or the primary outbreak cluster, though all were a part of the broad Pangolin lineage B.1.

**Figure 2A-D.**
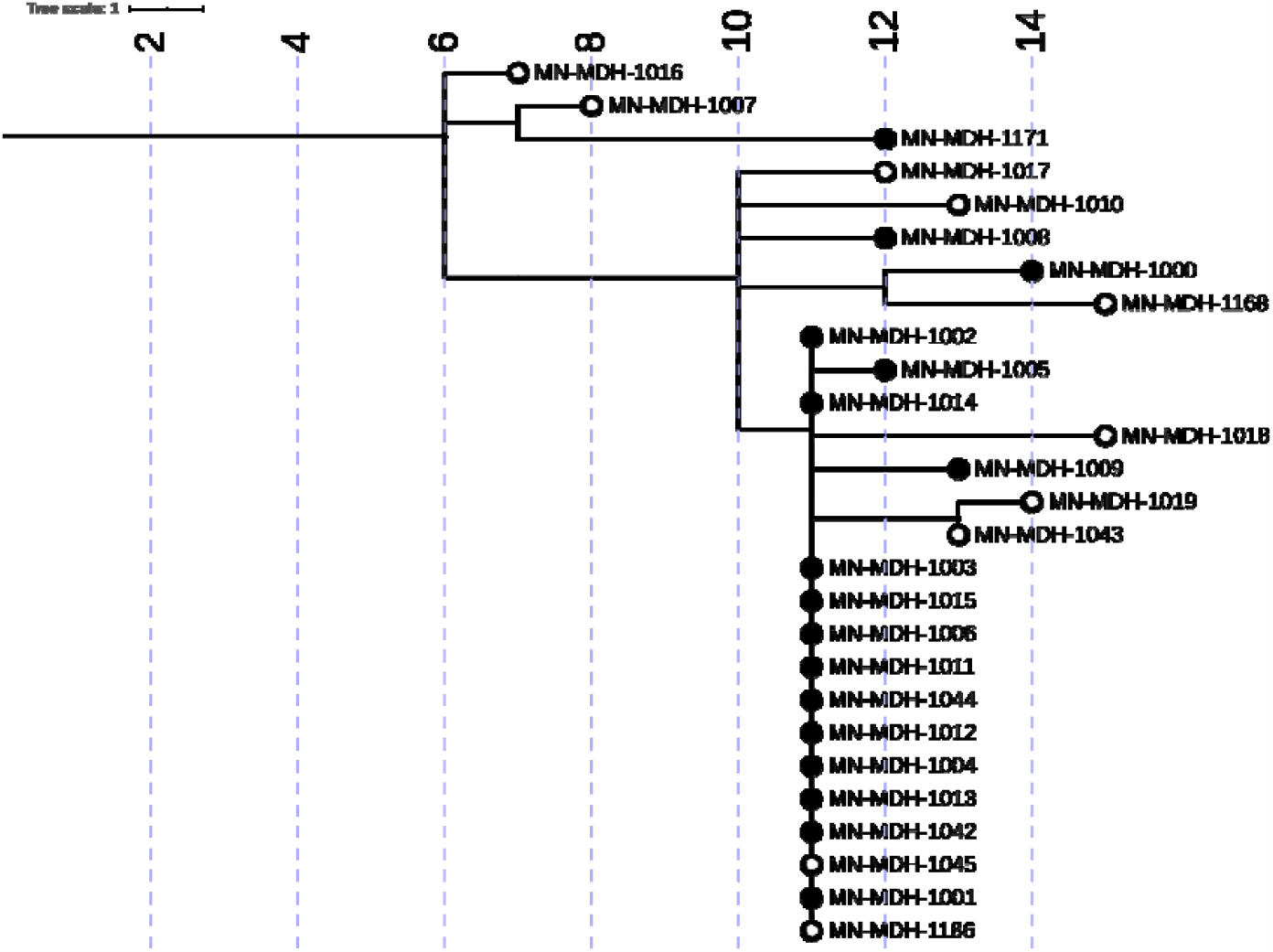
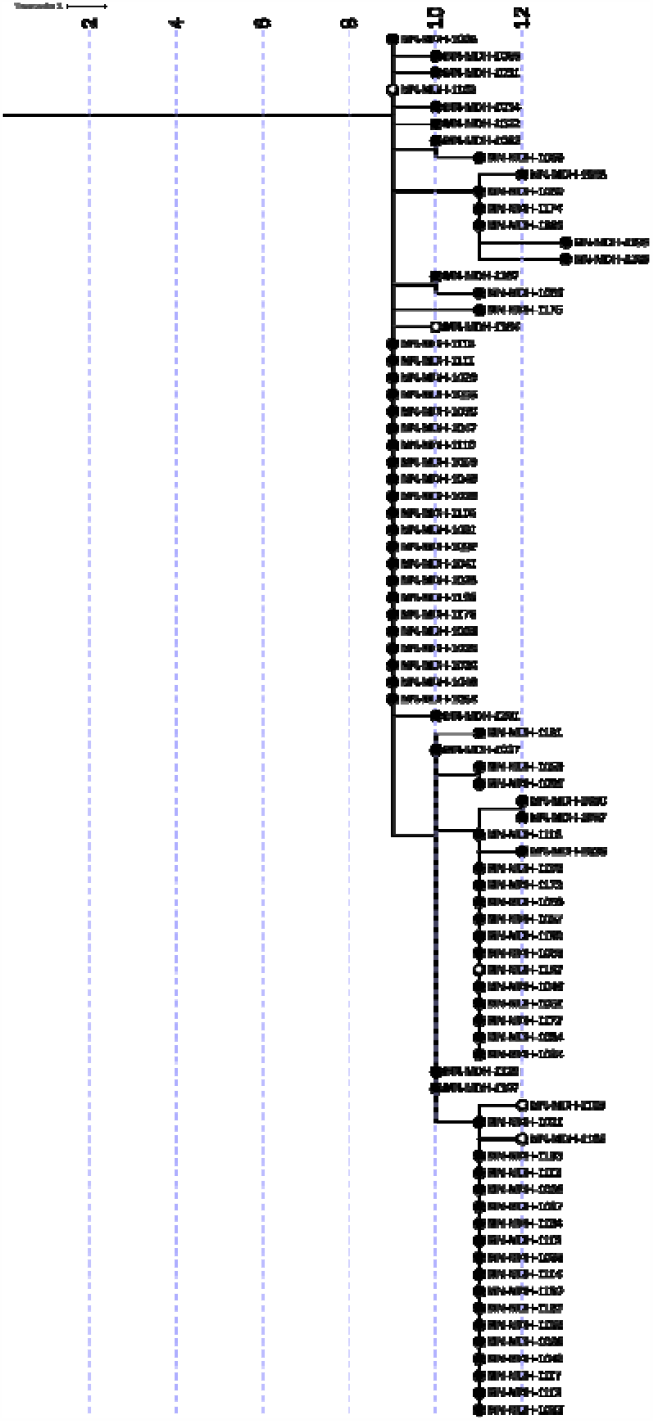
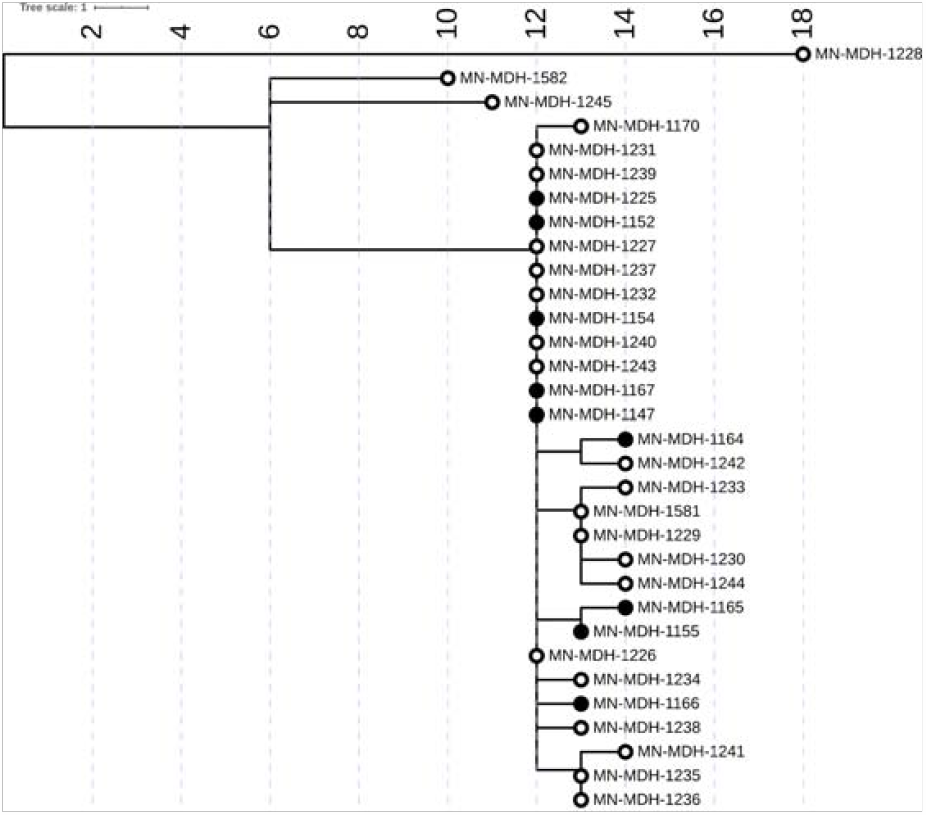
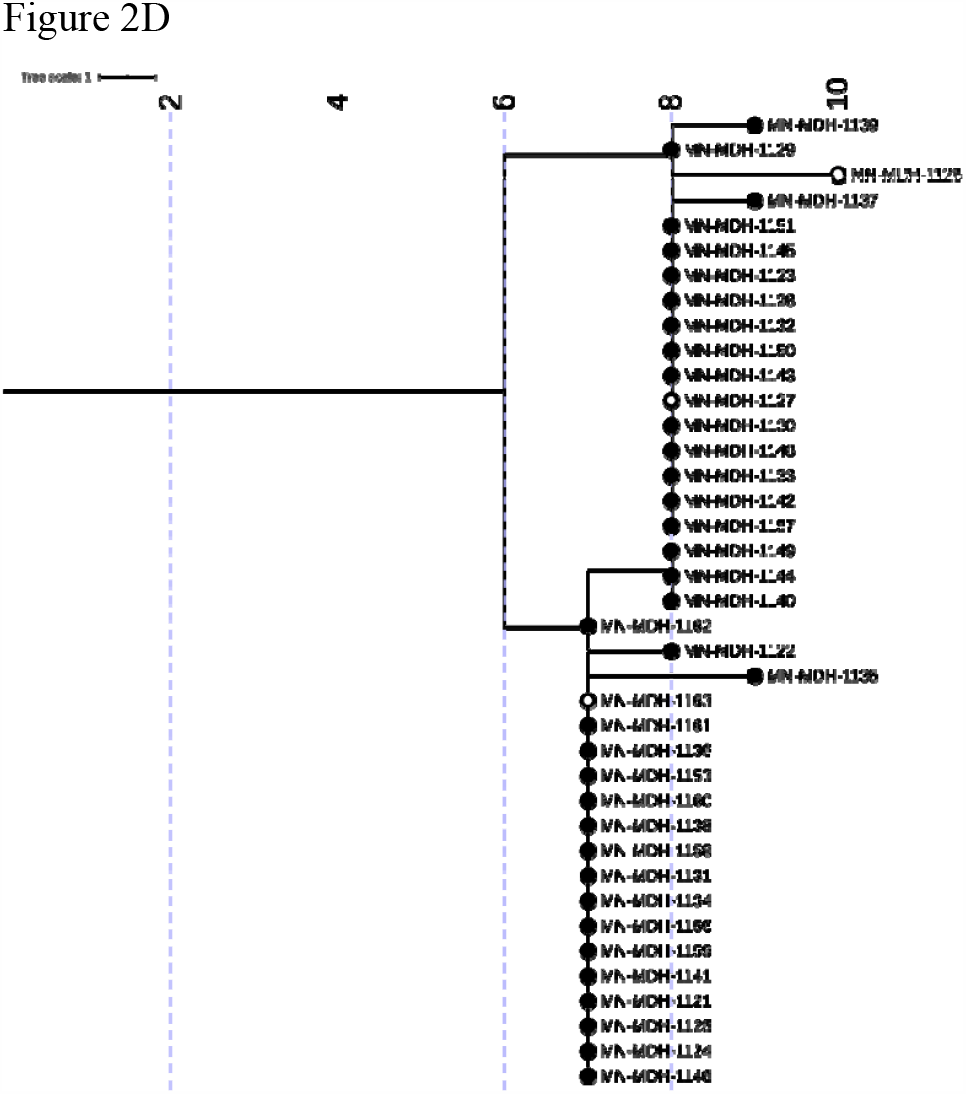
Phylogenetic tree of SARS-CoV-2 genome sequences associated with LTCF A from April 15 to June 11, 2020 (Figure 2A), LTCF B from April 29 to June 11, 2020 (Figure 2B), LTCF C from April 24 to June 30, 2020 (Figure 2C), and LTCF D from April 17 to May 15, 2020 (Figure 2D). Filled circles represent sequences taken from residents, open circles represent sequences from healthcare workers. IQ-TREE was used with the general time reversible substitution model for tree generation. Branch lengths were scaled to represent number of single nucleotide mutations as shown in the scale key.

In LTCF B from April 29 - June 11 (Figure 2B), 114 of 182 tested residents and 76 of 233 tested HCP were confirmed to be SARS-CoV-2 positive, following identification of a SARS-CoV-2 positive resident on April 29. All 82 sequenced specimens from this facility, including 77 residents (67.5% of cases) and 5 HCP (6.6% of cases), were closely related (Pangolin lineage B.1.116).

The first identified COVID-19 case at LTCF C (Figure 2C) was on April 24. Four positive HCP and 3 symptomatic residents were identified by April 30. Throughout May and June, facility-wide testing was implemented, with approximately 941 residents and staff tests performed, with 80 positive residents and 52 positive staff identified. Phylogenetic analysis of the 32 successfully sequenced genomes, including 9 residents (10.8% of cases) and 23 staff (41% of cases) showed 29 of the 32 cases were closely related (Pangolin lineage B.1.2). The remaining three cases (Pangolin lineages B.1 and B.4) were not closely related to each other nor identified with further transmission.

### Single Cluster in Correctional Facilities

In late March, 2020, an outbreak of SARS-CoV-2 was identified in Correctional Facility A (Figure 3A). The first identified case was an inmate who became symptomatic and tested positive for SARS-CoV-2 on March 25. By March 30^th^, a total of 7 confirmed cases and 6 suspected cases among the inmate population were identified. Between March 30 and April 7, 15 staff tested positive for SARS-CoV-2. Analysis of the genetic relatedness of the virus from 34 inmates (73.9% of cases) and 15 staff (18.3% of cases) from Correctional Facility A were all closely related (Pangolin lineage A.1).

**Figure 3A-B.**
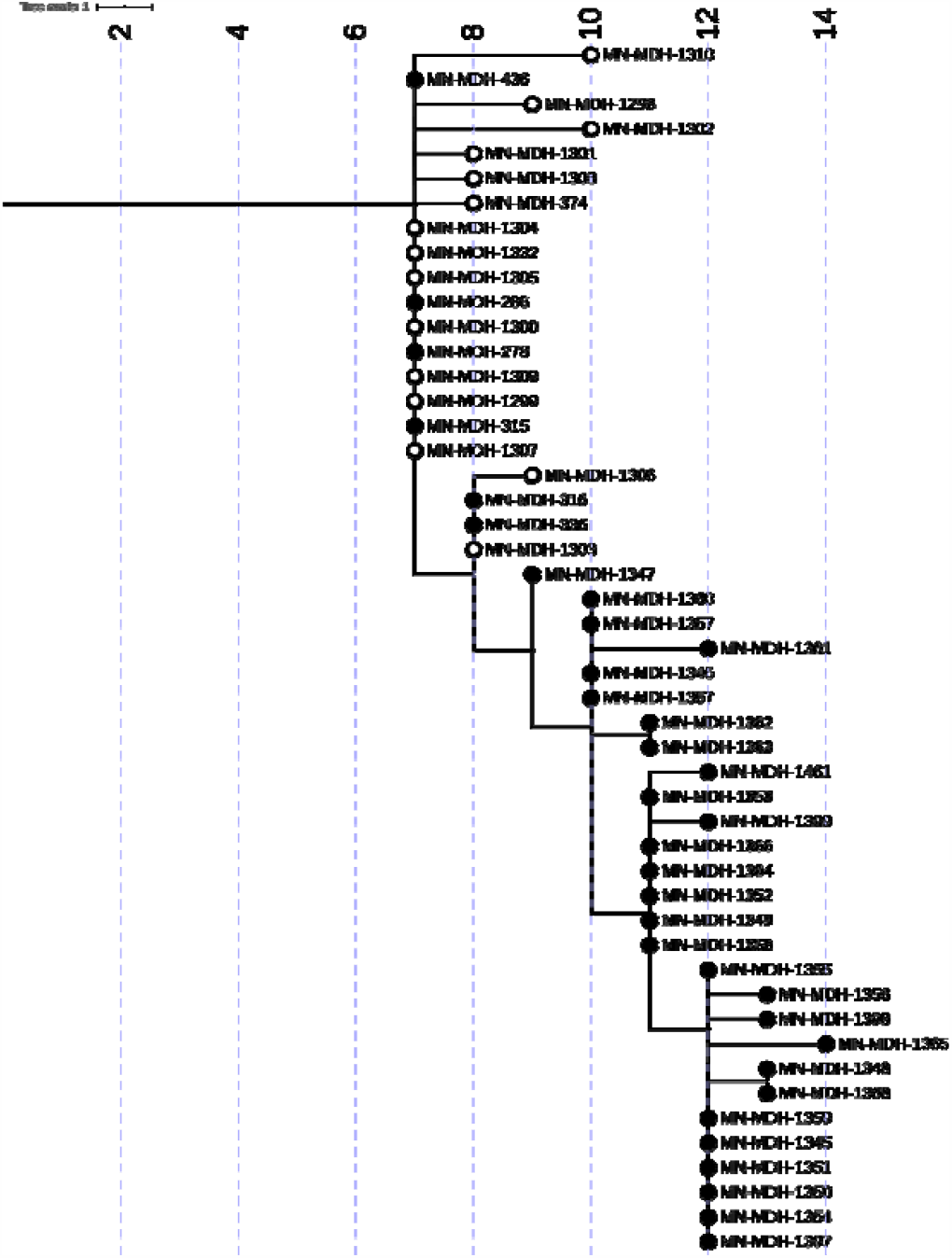
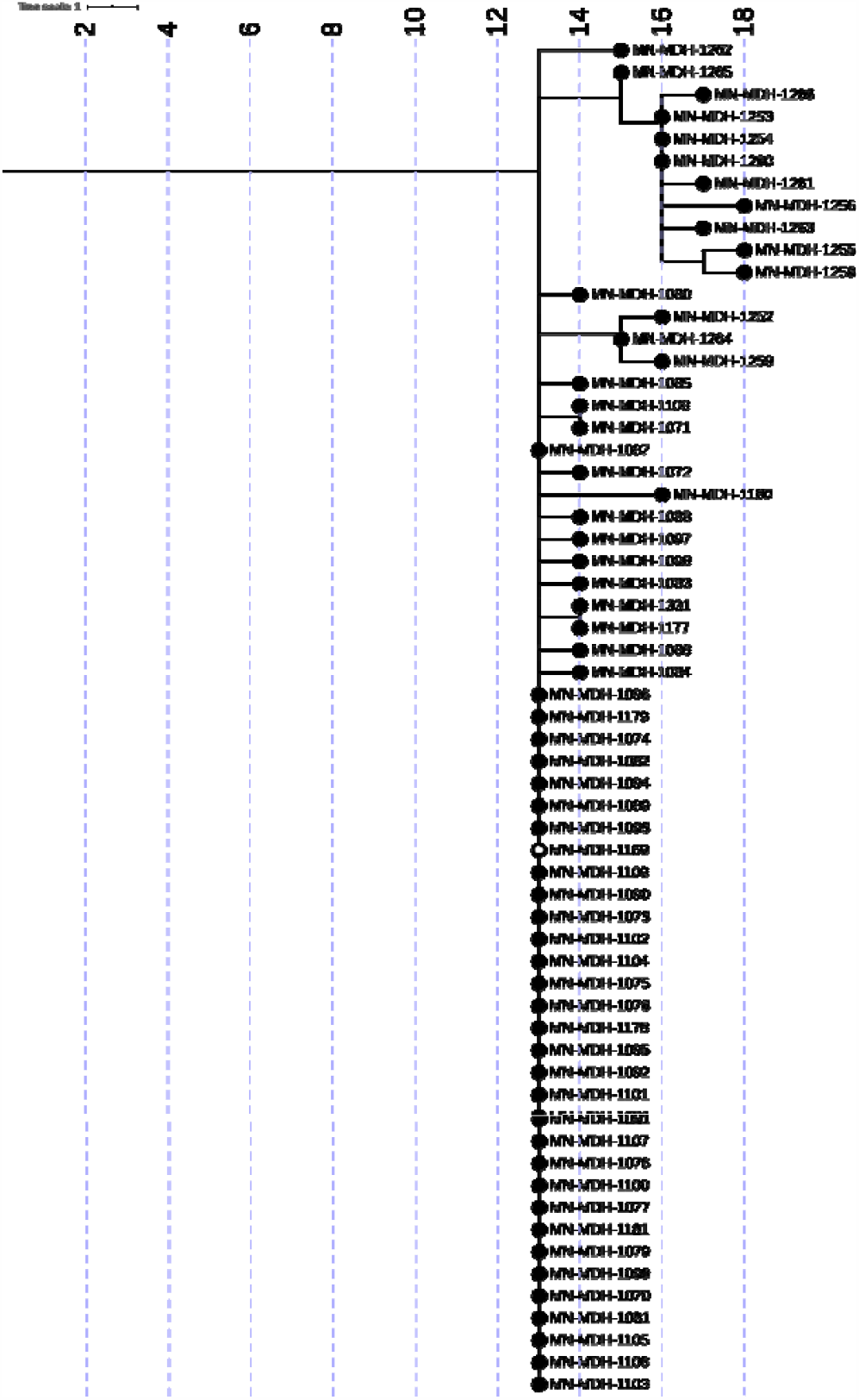
Phylogenetic tree of SARS-CoV-2 genome sequences associated with Correctional Facility A from March 25 to June 30, 2020 (Figure 3A), and Correctional Facility B from May 13 to June 30, 2020 (Figure 3B). Filled circles represent sequences taken from inmates, open circles represent sequences from facility staff. IQ-TREE was used with the general time reversible substitution model for tree generation. Branch lengths were scaled to represent number of single nucleotide mutations as shown in the scale key.

In early June, 2020, an outbreak was identified in Correctional Facility B (Figure 3B). The investigation revealed that an employee had developed symptoms consistent with COVID-19 on May 13, tested positive for SARS-CoV-2 on May 14, and was subsequently excluded from work and isolated at home. Approximately two weeks later, three additional cases, one staff member and two inmates from the same unit as the index patient, tested positive for SARS-CoV-A point-prevalence survey (PPS) on June 1 revealed 63 COVID-19 positive inmates among the 87 tested. Subsequent facility-wide testing of both staff and inmates identified cases in other units, 83 new cases in inmates and one new staff case identified among the approximately 2,200 persons tested. Phylogenetic analysis of this outbreak among the 1 staff member (0.5% of cases) and 60 inmates (29.1% of cases) at Correctional Facility B shows that all cases were closely related (Pangolin lineage B.1.2) and genetically identical to, or plausibly descended from, the sequence of SARS-CoV-2 from the index case.

### Linking LTCF C with Correctional Facility B

During the epidemiologic investigation at LTCF C, it was learned that a HCP at LTCF C was a household contact of a Correctional Facility B employee. Both individuals became symptomatic at the same time, and both subsequently tested positive in mid-May. SARS-CoV-2 genome sequences recovered from these two household contacts were identical to each other, and also to the genomic sequences recovered from 32 inmates at Correctional Facility B (Figure 4). In addition, this genomic sequence differs by only a single mutation (G5617T) from 13 cases sequenced from LTCF C.

**Figure 4.**
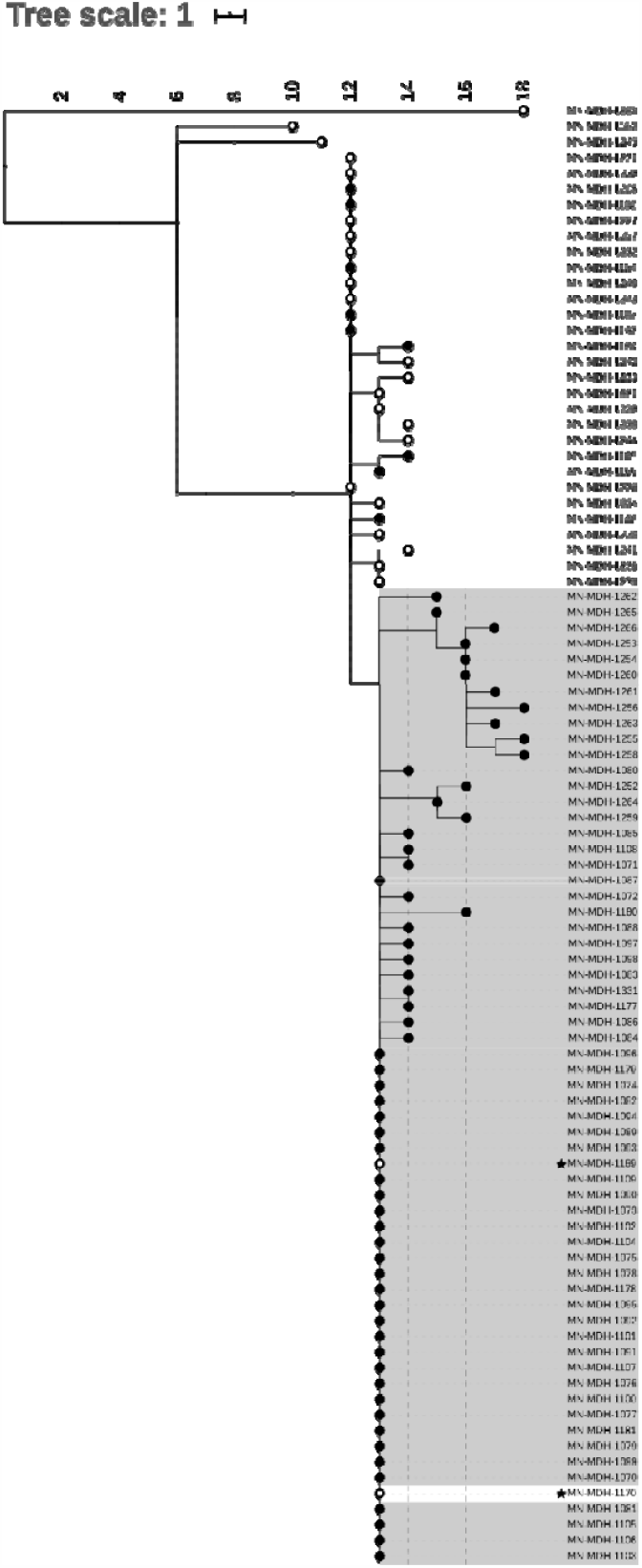
Phylogenetic tree of SARS-CoV-2 genome sequences associated with both LTCF C and Correctional Facility B. Filled circles represent sequences taken from inmates or residents, open circles represent sequences from facility staff or healthcare workers. Sequences from LTCF C are shown on a white background, and sequences from Correctional Facility B on a grey background. Sequences from two household contacts are noted with stars. IQ-TREE was used with the general time reversible substitution model for tree generation. Branch lengths were scaled to represent number of single nucleotide mutations as shown in the scale key.

### Two Distinct Clusters in a Long-Term Care Facility

LTCF D (Figure 2D) is a 100 bed facility with approximately 78 residents and 100 staff that experienced an outbreak beginning on April 17, 2020 with a symptomatic HCP. Identification of the first resident and staff case occurred on April 20, 2020, with subsequent testing identifying a total of 53 positive residents and 21 positive staff. While epidemiologically similar to other LTCF outbreaks, an analysis of the genetic relatedness among 39 sequenced cases demonstrated two distinct genetic clusters present in the facility during approximately the same time period. In contrast to the outbreaks in LTCFs A, B and C, viruses from both clusters appeared to circulate simultaneously throughout the facility, each contributing to the outbreak. All sequenced cases from LTCF D belonged to the broad Pangolin lineage B.1.

### Multiple Clusters in Meat Processing Plants

In early April 2020, an outbreak was detected at Processing Plant A (Figure 5A), a large primary and secondary meat processor. The outbreak at Processing Plant A continued for several weeks until mid-May when the number of cases among workers began to increase rapidly. From March 15 to July 1, there were 446 confirmed cases who reported working at Processing Plant A, including 4 (1%) cases who tested positive in March (management and office staff), 5 (1%) in April, 211 (47%) in May, and 226 (51%) in June. Analysis demonstrated that of the 16 samples (3.7% of cases) sequenced between March 15 and June 3, there were at least 6 unrelated clusters or singleton cases. While the majority of genomes sequenced from Processing Plant B belong to Pangolin lineages B.1, B1.2, B.1.26, one early case is genetically quite different (Pangolin lineage A.1). An interview confirmed that this early case had traveled out of the state during their exposure period (14 days before symptom onset).

**Figure 5A-B.**
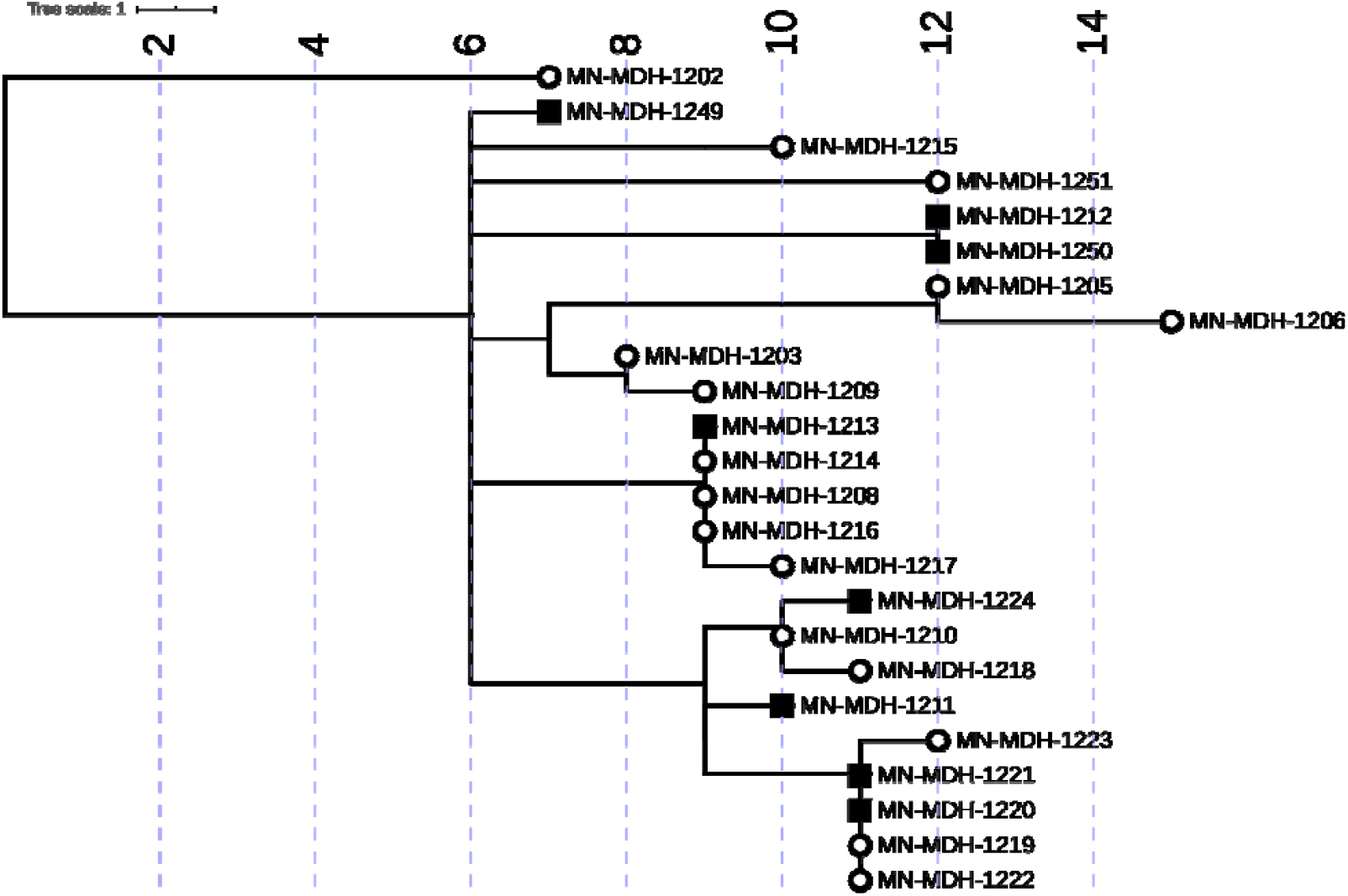
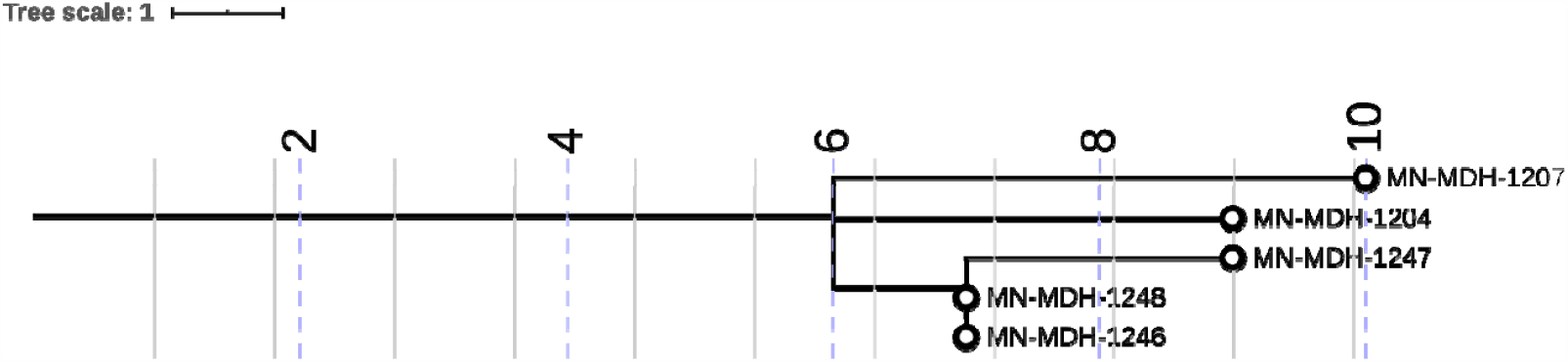
Phylogenetic tree of SARS-CoV 2 genome sequences associated with Processing Plant A and the surrounding community from March 15 to June 30, 2020 (Figure 5A), and Processing Plant B from April 11 to June 30, 2020 (Figure 5B). Open circles represent sequences from staff at Processing Plant A and squares represent sequences from the surrounding community. IQ-TREE was used with the general time reversible substitution model for tree generation. Branch lengths were scaled to represent number of single nucleotide mutations as shown in the scale key.

Between May 15 and June 1, we sequenced samples obtained from eight cases in the county where Processing Plant A is located was performed (Community Samples A). From these eight samples, a total of 5 clusters were identified. Five of the eight samples were closely related with three of the clusters from Processing Plant A, while the remaining three samples formed two distinct clusters. Four of the five sequences from Community Samples A that clustered with sequences from Processing Plant A had sequences that were identical to sequences from Processing Plant A, and all four had no known contact with a verified case.

In mid-April 2020, an outbreak was identified among employees at Processing Plant B (Figure 5B), another large meat processing plant. By May 1, there were 649 confirmed cases among workers at Processing Plant B. Sequencing of the 5 available samples from Processing Plant B (0.7% of cases) identified one cluster and two singleton genomes, all belonging to Pangolin lineage B.1.

## Discussion

WGS identified three primary patterns of genetic relatedness among cases in various outbreak settings: 1) outbreaks where cases were part of a single, genetically related cluster, 2) an outbreak with two unique clusters of cases, each contributing to the outbreak during the same time period, and 3) outbreaks where there were multiple genetically distinct sequences present.

Phylogenetic analyses of the viral sequences from available specimens associated with outbreaks in LTCF A, LTCF B and LTCF C were all consistent with at least one primary cluster impacting each facility, suggesting that a single introduction of SARS-CoV-2 into a facility can result in a widespread outbreak. This is similar to previous reports, where WGS has evidenced rapid spread in high density settings as opposed to multiple introductions contributing to the outbreak (*20*). Cases from LTCF D, in contrast, formed 2 distinct genetic clusters, one of 17 related samples and the other consisting of 22 samples. This is consistent with a potential scenario where there were two separate, independent introductions into the facility, and subsequent parallel intra-facility spread of each individually distinct sequence.

Phylogenetic analysis conducted in both LTCF A and LTCF C also demonstrated the presence of “outlier” SARS-CoV-2 viral sequences that were not genetically closely related to the primary cluster in each facility. This suggests community-acquired infection, and subsequent introduction of SARS-CoV-2 into the facility (*3*). Two of the three outlier cases at LTCF C tested positive over a month after the first identified case. Similarly, two of the three outlier cases identified at LTCF A were identified 10 days after the first identified case, while the third tested positive 28 days later. It is not possible to determine whether these introductions of distinct genetic sequences resulted in additional spread given that WGS characterization was not performed on all positive samples in each facility, and not all HCP or residents were tested.

However, the timing of the identification of these outlier cases after the date of the first identified primary case suggest that mitigation strategies implemented after the initial identification of the outbreak, including cohorting strategies, infection prevention and control measures (IPC), and correct use of personal protective equipment (PPE) may have been effective in preventing intra-facility transmission of these late outlier cases, as has been previously reported (*3, 22-23*).

WGS identified a different genetic landscape in processing plants, one in which several distinct sequences contributed to the facility outbreak. This finding is despite only 2.5% of positive cases at the processing plants having been sequenced, suggesting that increased sequencing may have identify even greater diversity. Additionally, several genomes identified at Processing Plant A were either identical or closely related to genomes present in the surrounding community (Community Samples A). Six of the eight sequenced Community Samples A were from persons with no known epidemiologic link to a case at Processing Plant A strongly suggesting an unrecognized connection. The benefit of WGS to identify previously unrecognized transmission patterns has previously been established (*20, 36*). While no definitive conclusions can be made regarding the direction of transmission, WGS provided strong evidence of a worker-community member spread, with communal housing, multigenerational families and group transportation as hypothesized factors potentially contributing to this transmission pattern.

WGS has shown to improve understanding of an outbreak upon retrospective analysis (*3, 20-22*), to provide justification for specific public health measures (*22-23*), and to add insight to transmission patterns in high-risk settings. Our work provides further support for the use of WGS in these situations, while identifying several additional public health implications. WGS has demonstrated that outbreaks in LTCFs and correctional facilities can be the result of a single introduction. Continued vigilance with facility-wide staff screening, and subsequent exclusion of symptomatic HCPs or staff and those with known or suspected contacts is imperative. WGS has demonstrated extensive intra-facility spread, with closely related sequences comprising all or the vast majority of cases contributing to the outbreak. Measures, such as IPC, consistent and correct use of PPE, cohorting of known positive residents and exclusion of positive HCPs must be maintained. WGS has also illuminated the transmission patterns in meat processing plants, including the multiple introductions identified through the multiple closely related sequences identified, and the related community strains. WGS has illustrated the need for community-level mitigation to prevent introductions in high-density worksites, including accessible community-wide testing, housing and transportation strategies, in addition to facility-level measures to prevent unintended introduction into the workplace.

There are several important limitations to this study. First, only a subset of specimens were available for sequencing due to different laboratory specimen retention policies. For example, at LTCF B, samples from only 5 staff were available for sequencing. Similarly, in Processing Plant B, only 5 samples were available due to a clinical testing laboratory protocol that resulted in the discarding of samples after approximately 7 days. In addition, not all available patient samples were able to be successfully sequenced, owing primarily to degraded or low concentrations of viral RNA in the specimen.

Another limitation is that not all staff and employees at the LTCFs, correctional facilities and processing plants agreed to be tested. Because of the incomplete genomic picture at each setting, definitive conclusions about single introductions in LTCF A and LTCF D are speculative, and these individual introductions may have actually resulted in some viral transmission that was not identified in the study.

Finally, we were not able to present sociodemographic data such as race or ethnicity pertaining to cases, or the facility population, associated with these eight outbreaks in Minnesota, due to limitations in the case investigation process and incomplete case data. This limitation is of particular importance, due to the disproportionate impact of COVID-19 on Black, Indigenous, and People of Color (BIPOC). Since BIPOC disproportionately experience incarceration, and a high proportion of meat processing plant employees are persons from immigrant communities, these settings can serve to amplify racial and ethnic health disparities related to COVID-19.

Long-term care facilities, correctional facilities, and high-density workplace settings have many factors that are hypothesized to contribute to rapid transmission of SARS-CoV-2. These include insufficient resources and training in IPC, difficulties implementing social distancing due to close habitation and/or work environment, and delayed case detection and access to care (*8, 11, 37*). WGS has demonstrated that many outbreaks in Minnesota were caused by single introductions of SARS-CoV-2, highlighting the importance of consistent and correct usage of PPE, rigorous and systematic IPC, environmental control measures, and systematic testing of residents and staff to identify asymptomatic cases. As this pandemic continues, community mitigation strategies and strong enforcement of policies to reduce the risk of introducing SARS-CoV-2 virus into congregate settings are more important than ever. Similarly, IPC and aggressive containment practices are vital to mitigate the spread of SARS-CoV-2 upon introduction into a facility. WGS can be a useful tool to supplement epidemiological information and examine the role of facility and community factors contributing to SARS-COV-2 outbreaks in high risk settings.

## Data Availability

There is no referred data.

## Acknowledgments

We would like to gratefully acknowledge the contributions from the following: MDH MEDSS Team, MDH Public Health Lab, MDH Case Investigation Team, MDH Outbreak Investigation Team, MDH Long-Term Care Facility Team, Matthew Binnicker and Joseph Yao at Mayo Clinic Laboratories, and Dr. Andrew C. Nelson and Sophia Yohe at M Health Fairview Molecular Diagnostics Laboratory.

## Disclaimers

The findings and conclusions in this report are those of the authors and do not necessarily represent the official position of the US Centers for Disease Control and Prevention.

## Author Bio

Dr. Nicholas Lehnertz is a Medical Specialist in Infectious Disease Epidemiology, Prevention and Control at the Minnesota State Department of Health. His current research involves the epidemiology of COVID-19 transmission patterns, clinical characteristics of pre-symptomatic COVID-19 infection in residents of long-term care facilities, and the understanding of human belief systems surrounding COVID-19.

Dr. Nicholas Lehnertz contact info: Minnesota Department of Health, 625 Robert Street North, St. Paul, MN 55164-0975 651-201-5270

## Footnotes

^1^ Only including individuals interviewed and where workplace information was provided.

^2^ An epidemiologic link among inmates is defined as residing on the same unit or ward within a 14-day time period of each other.

^3^ An epidemiologic link among correctional staff is defined as having the potential to have been within 6 feet for 15 minutes or longer while working in the facility during the 14 days prior to prior to the onset of symptoms; for example, worked on the same unit during the same shift. An epidemiologic link also requires that cases among correctional staff neither share a household together, nor were identified as close contacts with each other outside of the facility during the standard case investigation.

^4^ Facility-acquired COVID-19 infection in an inmate/resident is defined as a confirmed diagnosis 14 days or more after entry to the facility, without an exposure during the previous 14 days to another setting where an outbreak was known or suspected to be occurring.

^5^ Review consisted of discarding samples where read alignments to the Wuhan-Hu-1 reference had internal gaps greater than 125 nucleotides and removing trailing A nucleotides from consensus genomes in cases where genome alignments had reads mapped only to the reference sequence poly(A) tail and none of the preceding reference sequence.

^6^ No cases or samples sequenced after June 30 included in study. An outbreak is defined as closed if there are no new COVID-19 cases for 28 days after the onset date of the last case. The outbreak at Correctional Facility A was considered closed as of July 20, the outbreak at Correctional Facility B was considered closed as of August 5. The outbreaks at both Processing Plant A and Processing Plant B are considered ongoing as of November 6, 2020.

^7^ A full accounting of the outbreaks at LTCF A and LTCF B were previously published. *See* Taylor J, Carter RJ, Lehnertz N, et al. Serial Testing for SARS-CoV-2 and Virus Whole Genome Sequencing Inform Infection Risk at Two Skilled Nursing Facilities with COVID-19 Outbreaks — Minnesota, April–June 2020. MMWR Morb Mortal Wkly Rep 2020;69:1288–1295. DOI: http://dx.doi.org/10.15585/mmwr.mm6937a3

## Notes

### Competing Interest Statement

The authors have declared no competing interest.

### Funding Statement

Study was supported by the ELC Cares grant from CDC.

### Author Declarations

The manuscript was reviewed in accordance with standard CDC protocol, in which the approved CDC chain of command in the COVID 19 response division reviewed the manuscript and determined that it was non-research, public health response. As such, it was determined by CDC review to be exempt from further institutional review board evaluation. In summary, this manuscript and activity was reviewed by CDC and was conducted consistent with applicable federal law and CDC policy (see e.g., 45 C.F.R. part 46, 21 C.F.R. part 56; 42 U.S.C. 241(d); 5 U.S.C 552a; 44 U.S.C. 351 et seq.).

